# Variations in the Circle of Willis in a large population sample using 3D TOF angiography: The Tromsø Study

**DOI:** 10.1101/2020.06.15.20129353

**Authors:** Lars B. Hindenes, Asta K. Håberg, Liv Hege Johnsen, Ellisiv B. Mathiesen, David Robben, Torgil R. Vangberg

## Abstract

The main arteries that supply blood to the brain originate from the Circle of Willis (CoW). The CoW exhibit considerable anatomical variations which may have clinical importance, but the variability is insufficiently characterised in the general population. We assessed the anatomical variability of CoW variants in a community-dwelling sample (N = 1,864, 874 men, mean age = 65.4, range 40 – 87 years), and independent and conditional frequencies of the CoW’s artery segments. CoW segments were classified as present or missing/hypoplastic (w/1mm diameter threshold) on 3T time-of-flight magnetic resonance angiography images. We also examined whether age and sex were associated with CoW variants. We identified 47 unique CoW variants, of which five variants constituted 68.5% of the sample. The complete variant was found in 11.9% of the subjects, and the most common variant (27.8%) was missing both posterior communicating arteries. Conditional frequencies showed patterns of interdependence across most missing segments in the CoW. CoW variants were associated with mean-split age (P = .0147), and there was a trend showing more missing segments with increasing age. We found no association with sex (P = .0526). Our population study reinforced age as associated with CoW variants, suggesting reduced collateral supply with older age.

## Introduction

The primary blood supply to the brain originate from three arteries, the left and right internal carotid artery and the basilar artery. These three arteries anastomose (i.e. join) to form the Circle of Willis (CoW) at the base of the brain (Supplementary Figure S1). The circular arrangement of arteries in the CoW enables redistribution of blood flow when arteries in, or upstream of the CoW experience reduced flow capacity. This so-called collateral ability of the CoW ensures redundancy in the blood supply to the brain. Surprisingly, segments in the CoW are commonly missing or hypoplastic (i.e. underdevelopment of segment diameters), rendering the CoW incomplete, and consequently reducing the collateral ability of the CoW and increasing the brain’s vulnerability to changes in the blood flow (1–3). Earlier studies have shown that increasing age and male sex are associated with incomplete CoW variants (1,4). The genetic influence on CoW variants remains inconclusive, with one study showing a hereditary effect for incomplete CoW variants (5), while a twin study failed to find any effect (6).

The anatomy of the CoW has clinical relevance since incomplete CoW variants are associated with an increased risk of cerebrovascular disease. Studies on patient samples find that incomplete CoW variants are associated with stroke (7,8), aneurisms (9,10) and white matter hyperintensities (11–14). The collateral ability of CoW variants are also important in certain surgical procedures (15,16). However, well-powered studies have not been performed using population-based samples, and therefore it is not clear whether incomplete variants pose a risk for cerebrovascular disease in the general population.

Our understanding of the anatomical variability in the CoW is limited by greatly varying prevalence estimates. For example, the estimated prevalence of the complete variant range from 12.2% (3) to 45.0% (6). Differences in sample characteristics (1,5,15,17), sample size (18), and measuring techniques (1,17) (i.e. dissection, CT or MRI) are assumed sources of variability in these estimates. Additionally, many CoW classification schemes are impossible to fully compare, complicating comparison across studies (1,14,19–21). Common classification schemes based on diameters include different approaches such as using diameter thresholds for arteries (8,15,19,20,22,23), comparing diameters of arteries relative to other arteries’ diameter (3,9,24), or a mix of both (1,11,17,25). Other schemes include splitting the classification into the anterior and posterior circulation of the CoW (1,8). or not distinguishing between left and right sided variants (2,3,21). One study did not even report a classification scheme (2).

The primary goal of this study was to report population-based estimates of the prevalence of CoW variants based on high resolution 3T MR angiography images using a more rigorous and reproducible classification scheme compared to previous studies. We also examined whether CoW variants were associated with age and sex. Lastly, to provide a richer description of the anatomical variability of the CoW we estimated the frequency of individual missing arterial segments in the CoW and their conditional frequencies, independently of CoW variants.

## Materials and methods

### The Tromsø Study

The Tromsø Study is a population-based cohort study recruiting from the Tromsø municipality in Norway. This study has been performed every six to seven years since 1974 and the seventh survey (Tromsø 7) finished in 2015 – 2016. Tromsø 7 consisted of two visits. All inhabitants above age 40 were invited to the 1^st^ visit, and 20,183 subjects participated (65% participation rate). A subset of participants in the first part of the Tromsø 7 Study were invited to a 2^nd^ visit, where 8,346 subjects participated. Of these, 2,973 were invited to partake in a cross-sectional magnetic resonance (MR) study. Of the invited, 525 declined, 396 did not respond, 169 had conditions prohibiting MR examinations, and five had moved or were dead. Furthermore, for 14 cases we were unable to find at least one of three baseline MRI series, consequently yielding 1,864 subjects with time-of-flight (TOF) angiography series, T1-weighted series, and T2-weighted fluid-attenuated inversion recovery (FLAIR) series (see Supplementary Figure S2 for overview of the inclusion process). The study was approved by the Regional Committee of Medical and Health Research Ethics Northern Norway (2014/1665/REK-Nord) and carried out in accordance with relevant guidelines and regulations at UiT The Arctic University of Norway. All participants gave written informed consent before participating in the study.

### MRI protocol

Participants were scanned at the University Hospital North Norway in a 3T Siemens Skyra MR scanner (Siemens Healthcare, Erlangen, Germany). A 64-channel head coil was used in most examinations, but in 39 examinations, a slightly larger 20-channel head coil had to be used. The MRI protocol consisted of a 3D T1-weighted series, a 3D T2-weighted FLAIR series, a susceptibility weighted series and a TOF angiography series, with a total scan time of 22 minutes. Only the TOF images were used in this study. These were acquired with a 3D transversal fast low angle shot sequence with flow compensation (TR/TE = 21/3.43 ms, parallel imaging acceleration factor 3, FOV 200 × 181mm, slice thickness 0.5 mm, 7 slabs with 40 slices each). Reconstructed image resolution was 0.3 × 0.3 × 0.5mm. The slice prescription was automatically aligned to a standardized brain atlas ensuring consistency across examinations (26).

### Classification of CoW variants

TOF images were evaluated by LBH, using a program created in MeVisLab (v3.0.1). The program displays the TOF images both as a 3D rendering or a maximum intensity projection (MIP), and in 2D with a lumen diameter measurement tool. For rating an artery as present, the following criteria were used: (1) visible along its entire segment on the 3D rendering, (2) have a diameter larger than 1 mm, (3) connected to other arteries as in the complete textbook CoW. We did not differentiate between missing and hypoplastic segments due to image resolution. Therefore, missing and hypoplastic cases were referred to as missing. The classification criteria are illustrated in Figure 1 with different degrees of hypoplastic/missing arteries. Compare it to Supplementary Figure S1 for a complete “textbook-type” CoW.

**Figure 1.**
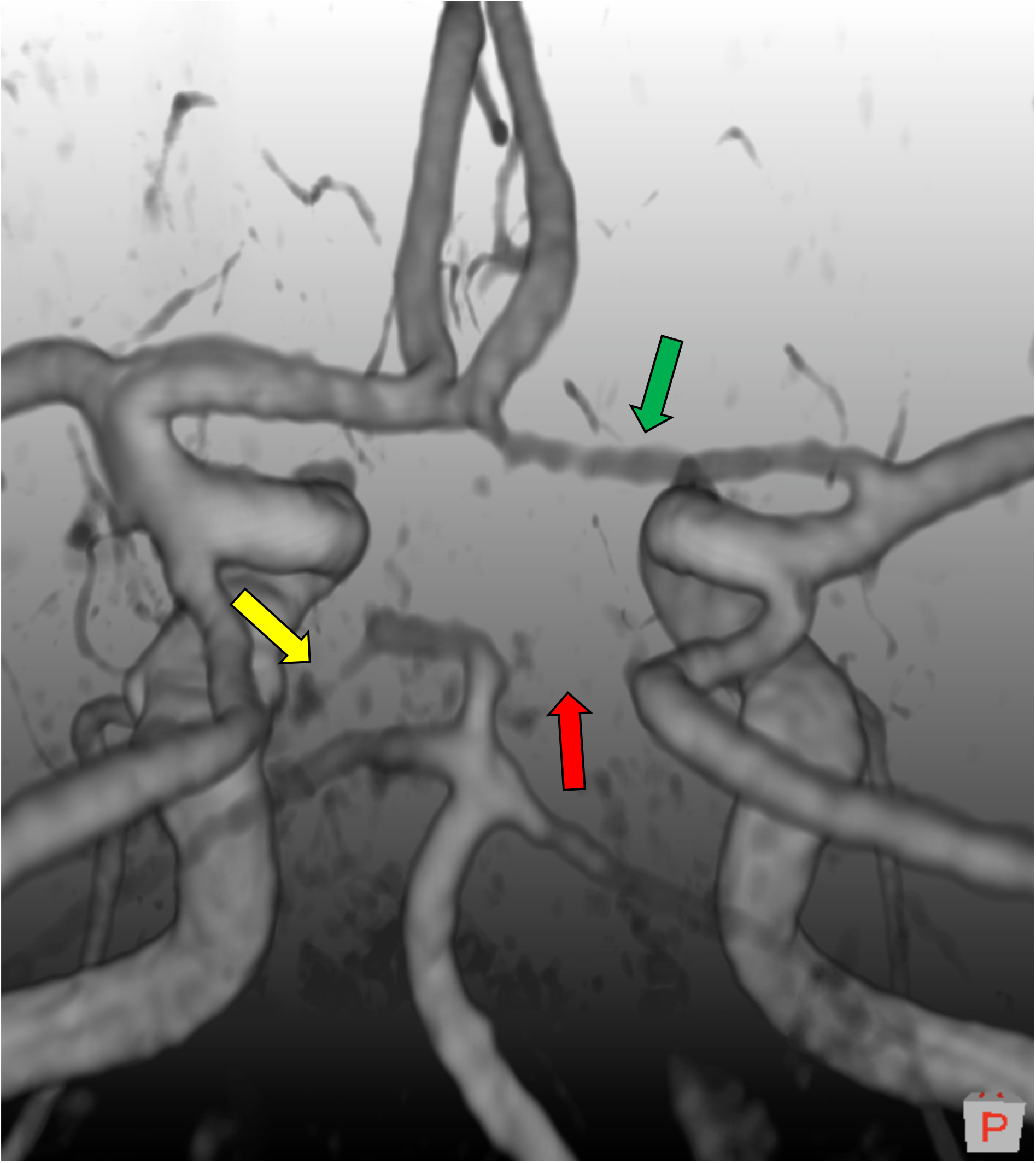
A 3D volume rendering of a time-of-flight magnetic resonance image depicting three classification scenarios we can encounter with our classification scheme. Green arrow: The right anterior cerebral artery is present. Yellow arrow: The left posterior cerebral artery is hypoplastic or missing, and just below 1mm in diameter. Red arrow: The right posterior cerebral artery is clearly missing. The configuration itself is of bilateral missing posterior cerebral artery (2P) type. Image follows neurological convention, where left is left and right is right. An orientation cube in the lower right corner show orientation, and its P denotes posterior.

The arteries of interest were the interconnecting arteries in the CoW, and the three largest in-flow arteries (Supplementary Figure S1). These included the internal carotid artery (ICA), middle cerebral artery (MCA), proximal anterior cerebral artery (ACA), anterior communicating artery (ACoA), posterior communicating artery (PCoA), proximal posterior cerebral artery (PCA) and vertebrobasilar artery (VBA). Consequently, the persistent primitive trigeminal artery and other special arteries as described in Dimmick et al. (27), were ignored. Care was taken to distinguish PCoAs from anterior choroidal arteries and PCAs from superior cerebellar arteries. Arterial segments that did not connect to their expected locations were classified according to the third criterion. For instance, variations in the ACoA, of which there are many of (3), were all regarded as a single ACoA. Furthermore, a posterior CoW variant named unilateral dual PCA (see Supplementary Figure S3) or hyperplastic anterior choroidal artery (27) was categorized as missing PCoA using the third criterion, because of the missing connection between the PCoA and its ipsilateral PCA. This simplification via the third criterion does not compromise the descriptions of the collateral flow ability within each CoW variant.

We labelled the CoW variants using a nomenclature similar to previous studies (1,9), where each variant’s name signified the missing segments. For brevity, ACA were denoted by “A”, ACoA by “Ac”, PCA by “P”, PCoA by “Pc”, ICA by “I”, MCA by “M”, and VBA by “V”. Alternatively, when no artery segment was missing a complete CoW was denoted by “O”. To specify whether a missing segment was in the left or right hemisphere, an “l” or “r” suffix is used. If the same segment was missing on both sides the number “2” was instead used as a prefix, e.g. “2Pc” for the variant where both PCoA are missing. This scheme ensured unique names for all CoW variants. See Figure 2 for illustrations of variants with their corresponding label.

**Figure 2.**
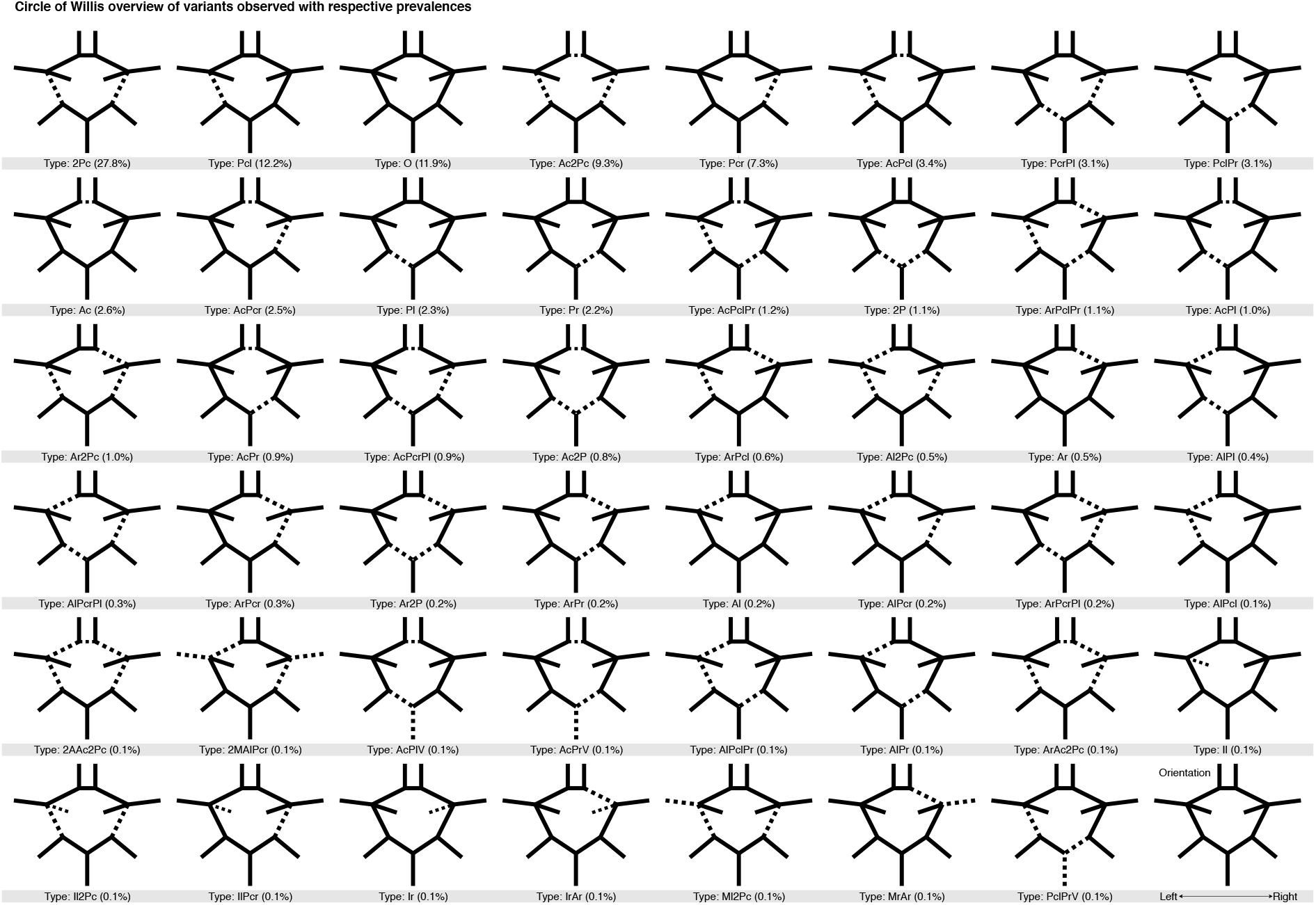
A full graphical overview of all Circle of Willis variants observed and subsequently classified in the current study, and with corresponding frequency. All variants are sorted first by (descending) frequency and then, in case of equal frequencies, by alphanumerical ordering. Each variant’s name is put together by the missing segments with the following notation: O = Complete variant (no missing arteries), Ac = Anterior communicating artery, A = Anterior cerebral artery, Pc = Posterior communicating artery, P = Posterior cerebral artery, I = Internal carotid artery, M = Middle cerebral artery, V = Vertebrobasilar artery, while the suffixes “r” and “l” denote right and left lateralization of arteries. The prefix “2” denote bilateral missing arteries.

A random sample (N = 100) was blinded and reclassified by the same rater (LBH), and also another rater (TRV) blinded to the original classification, in order to measure intra- and inter rater accuracy.

### Comparison with other studies

To contextualise our CoW variant frequencies, we wanted to compare with other studies. Unfortunately, to our knowledge, there is only one other CoW TOF MR study with a larger sample size than ours. The study is in 2246 healthy Chinese men (3), and we were able to perform comparisons with their study with only minor adaptations of the classification of the CoW variants. Main changes included removing left and right lateralization in our CoW variants, and translating their CoW variants to our nomenclature. Further information about the comparison is found in the Supplementary Data and the Supplementary Spreadsheet.

### Statistical analysis

We split the CoW data at mean age of participants, grouping subjects into a “younger” and “older” group (Table 1). We also grouped the subjects into age per decade (5 categories, 40 years to 90 years). Formulas used to calculate all frequencies for every CoW segment and variant are described in the Supplementary Methods. CoW variants observed less than ten times were grouped into a single composite category of rare variants (Supplementary Table S1). This composite category was created to include all subjects when testing without having too few observations in the array elements (see Supplementary Methods for details). The Cochran-Mantel-Haenszel test was used to test whether CoW variant frequencies were associated with sex, and age. This test allows for testing conditional independence between two factors while controlling for a third factor. As such, we used the Cochran-Mantel-Haenszel test to test for conditional independence between CoW variants and sex while controlling for the dichotomic age variable, and to test for conditional independence between CoW variants and dichotomic age while controlling for sex. Both of these tests had array dimensions 23 × 2 × 2. The effect of age was further examined by plotting the distribution of CoW variants for each decade. To assess whether sex might affect this plot, a 5 × 2Chi-squared test between age per decade and sex was performed to assess independence (Table 1). We considered a Bonferroni corrected P < 0.05 as significant (nominal P < 0.0167). At last, the accuracy metric was used to assess the intra- and inter rater validation. All computations were performed in R (v3.4.4) and three figures were created using the ggplot2 package (28).

**Table 1.** Age distributions per sex of subjects.

|  | **Men n = 874:** | **Women n = 990:** |
| --- | --- | --- |
| **Mean split age** |  |  |
| Above mean age* | 503 (57.6) | 513 (51.8) |
| Below mean age* | 371 (42.4) | 477 (48.2) |
| **Age by decade** |  |  |
| 40 - 49 | 84 (9.6) | 123 (12.4) |
| 50 - 59 | 135 (15.4) | 177 (17.9) |
| 60 - 69 | 313 (35.8) | 353 (35.6) |
| 70 - 79 | 273 (31.2) | 264 (26.7) |
| 80 - 89 | 69 (7.8) | 73 (7.4) |

(\*) = Average age is 65.4 years.

## Results

### Study participants

The mean age for all participants was 65.4 years (SD = 10.6). There were 874 men (47%), mean age 66.1 years (SD = 10.4, range = 40 – 86 years), and 990 women (53%), mean age 64.7 years (SD = 10.7, range = 41 – 87 years). Distributions of both mean-split age and decade age groupings with respect to sex are shown in Table 1, and distribution of age for men and women are shown in Supplementary Figure S4.

### Frequencies of CoW variants

We found 47 unique variants of the CoW (Figure 2). Of these, 22 made up 96.8% of the sample (Table 2), while the remaining 25 variants had less than ten observations each and constituted in total only 3.2% of the sample (Supplementary Table S1). The most common variants were, 2Pc (27.8%), with both PCoA segments missing, Pcl (12.2%), with the left PCoA segment missing, the complete O variant (11.9%), Ac2Pc (9.3%), with the ACoA and both PCoA missing, and Pcr (7.3%) with the right PCoA missing. These five most common CoW variants constituted 68.5% of the total sample. These findings suggest that only about 12% of the adult population have a complete CoW, while the remaining 88% have one or more missing segments, thus reducing their collateral capacity.

**Table 2.** Frequencies of common Circle of Willis variants for the whole sample, and their frequencies for men and women, and for being below and above mean age [Number of cases (Percentage of column total)].

| **Variant:** | **Total**<br>**n = 1,864:** | **Men**<br>**n = 874:** | **Women**<br>**n = 990:** | **Below**<br>**mean age**<br>**n = 848:** | **Above**<br>**mean age**<br>**n = 1,016:** |
| --- | --- | --- | --- | --- | --- |
| **2Pc** | 518 (27.8) | 266 (30.4) | 252 (25.5) | 222 (26.2) | 296 (29.1) |
| **Pcl** | 227 (12.2) | 95 (10.9) | 132 (13.3) | 116 (13.7) | 111 (10.9) |
| **O** | 221 (11.9) | 88 (10.1) | 133 (13.4) | 123 (14.5) | 98 (9.6) |
| **Ac2Pc** | 173 (9.3) | 72 (8.2) | 101 (10.2) | 73 (8.6) | 100 (9.8) |
| **Pcr** | 137 (7.3) | 69 (7.9) | 68 (6.9) | 71 (8.4) | 66 (6.5) |
| **AcPcl** | 63 (3.4) | 33 (3.8) | 30 (3.0) | 31 (3.7) | 32 (3.1) |
| **PcrPl** | 58 (3.1) | 26 (3.0) | 32 (3.2) | 29 (3.4) | 29 (2.9) |
| **PclPr** | 57 (3.1) | 35 (4.0) | 22 (2.2) | 25 (2.9) | 32 (3.1) |
| **Ac** | 49 (2.6) | 19 (2.2) | 30 (3.0) | 26 (3.1) | 23 (2.3) |
| **AcPcr** | 46 (2.5) | 18 (2.1) | 28 (2.8) | 20 (2.4) | 26 (2.6) |
| **Pl** | 43 (2.3) | 16 (1.8) | 27 (2.7) | 13 (1.5) | 30 (3.0) |
| **Pr** | 41 (2.2) | 22 (2.5) | 19 (1.9) | 17 (2.0) | 24 (2.4) |
| **AcPclPr** | 23 (1.2) | 11 (1.3) | 12 (1.2) | 9 (1.1) | 14 (1.4) |
| **2P** | 21 (1.1) | 11 (1.3) | 10 (1.0) | 9 (1.1) | 12 (1.2) |
| **ArPclPr** | 21 (1.1) | 9 (1.0) | 12 (1.2) | 7 (0.8) | 14 (1.4) |
| **AcPl** | 19 (1.0) | 10 (1.1) | 9 (0.9) | 8 (0.9) | 11 (1.1) |
| **Ar2Pc** | 19 (1.0) | 10 (1.1) | 9 (0.9) | 9 (1.1) | 10 (1.0) |
| **AcPr** | 17 (0.9) | 6 (0.7) | 11 (1.1) | 9 (1.1) | 8 (0.8) |
| **AcPcrPl** | 16 (0.9) | 8 (0.9) | 8 (0.8) | 3 (0.4) | 13 (1.3) |
| **Ac2P** | 14 (0.8) | 7 (0.8) | 7 (0.7) | 3 (0.4) | 11 (1.1) |
| **ArPcl** | 11 (0.6) | 2 (0.2) | 9 (0.9) | 4 (0.5) | 7 (0.7) |
| **Al2Pc** | 10 (0.5) | 4 (0.5) | 6 (0.6) | 3 (0.4) | 7 (0.7) |
| **Rare/**<br>**Other** | 60 (3.2) | 37 (4.2) | 23 (2.3) | 18 (2.1) | 42 (4.1) |

### Comparison of adapted CoW estimates with a previous study

After adapting the CoW estimates, we were able to compare 18 of our resulting prevalence estimates to the well-powered Chinese study (3) (Supplementary Data and Supplementary Spreadsheet). This comparison showed excellent agreement; with mean and median percent point differences of 1.6 and 0.8 respectively (range 0.1 - 10.6 percent points). The difference between the complete variant estimates was only 0.3 percent points. Most of the total 28.4 percent point difference could be attributed to variants missing PCoA or PCA. In the end, we compared 99.2% of our sample with 100% of the other study, resulting in only an additional 0.8 percent point bias. In sum, the comparison showed near-perfect agreement for nearly all CoW variants.

### Frequencies of missing segments independent of CoW variant

The frequencies of missing CoW segments in the whole sample are shown in Figure 3. The right and left PCoA were most frequently missing (60.6% and 53.6%), followed by the ACoA (22.7%). There was a notable right-left asymmetry in the frequencies of missing ACA and PCoA, but not for PCA. The right-to-left ratio for ACAs (4.3%/1.8%) was large, considering how infrequent the ACAs were missing.

**Figure 3.**
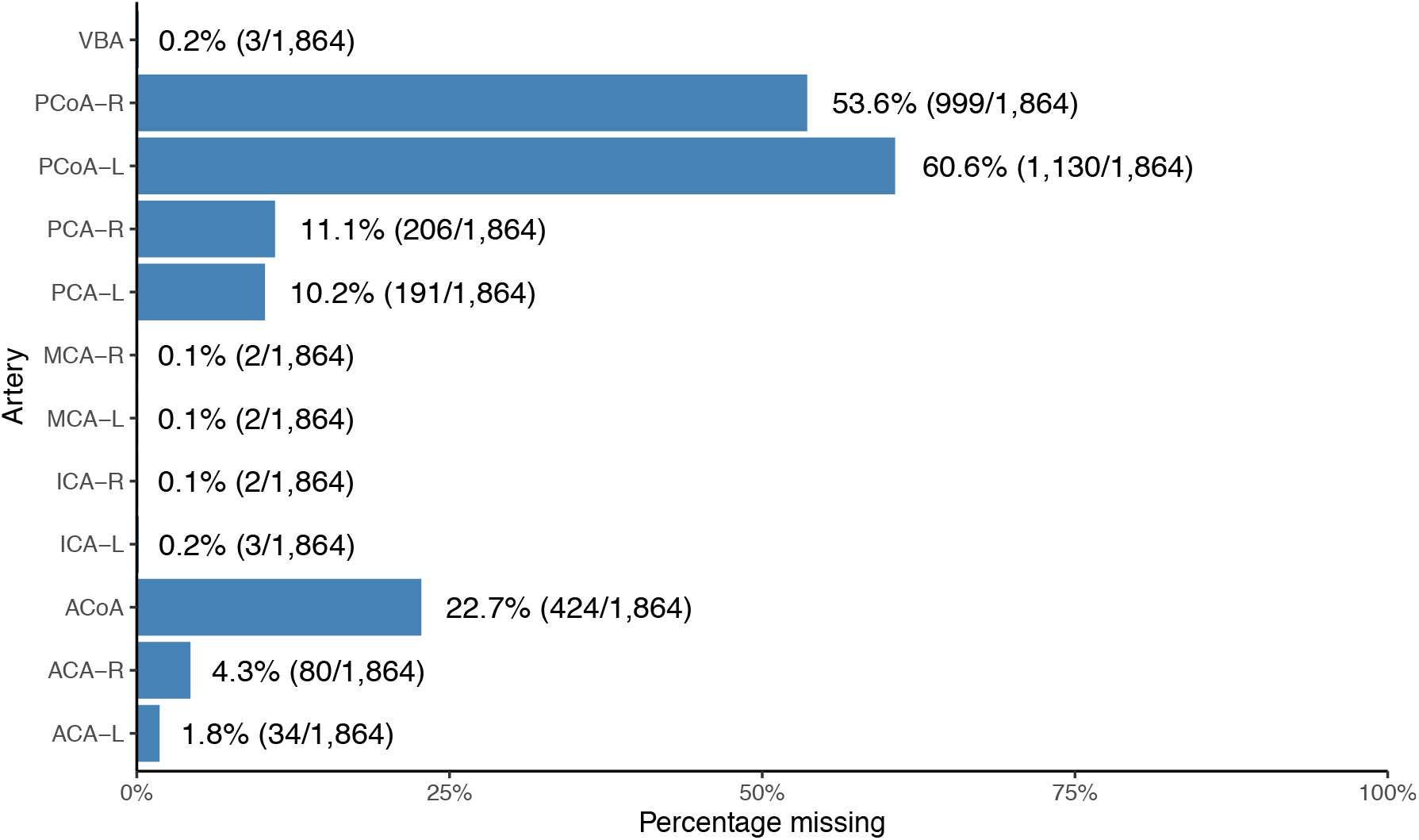
Frequency at which each artery is missing in the Circle of Willis in the whole sample, independently of other arteries and variants. Nominators and denominators are in corresponding parentheses, and represent respectively the number of times an artery is missing and the total number of subjects. ACA: Anterior cerebral artery. ACoA: Anterior communicating artery. PCoA: Posterior communicating artery. PCA: Posterior cerebral artery. ICA: Internal carotid artery. MCA: Middle cerebral Artery. VBA: Vertebrobasilar artery. Hemispheric left and right lateralization are denoted by “L” and “R” respectively.

### Pairwise conditional frequencies of missing segments

The heatmap of conditional frequencies (Figure 4) shows the conditional probabilities between CoW segments that were commonly missing, i.e. PCoA, PCA, ACA and ACoA (Figure 3). Although the conditional frequencies ultimately reflect the observed variant frequencies, the heatmap representation reveals several interesting patterns. First, ACoA was seldom missing if the left or right ACA was missing. Second, the ACA, PCoA and PCA segment pairs had approximately equal probability of being missing if the ACoA was missing. Third, if ACA was missing on one side, it was much more likely that the PCA was missing on the same side than on the opposite side, suggesting an ipsilateral pattern. Fourth, a contralateral pattern existed between ACA and PCoA, i.e., if the ACA was missing on one side, it was more likely that the PCoA was missing on the other side. Lastly, similar contralateral patterns were also seen between PCA and PCoA, the PCoA pair, and the PCA pair.

**Figure 4.**
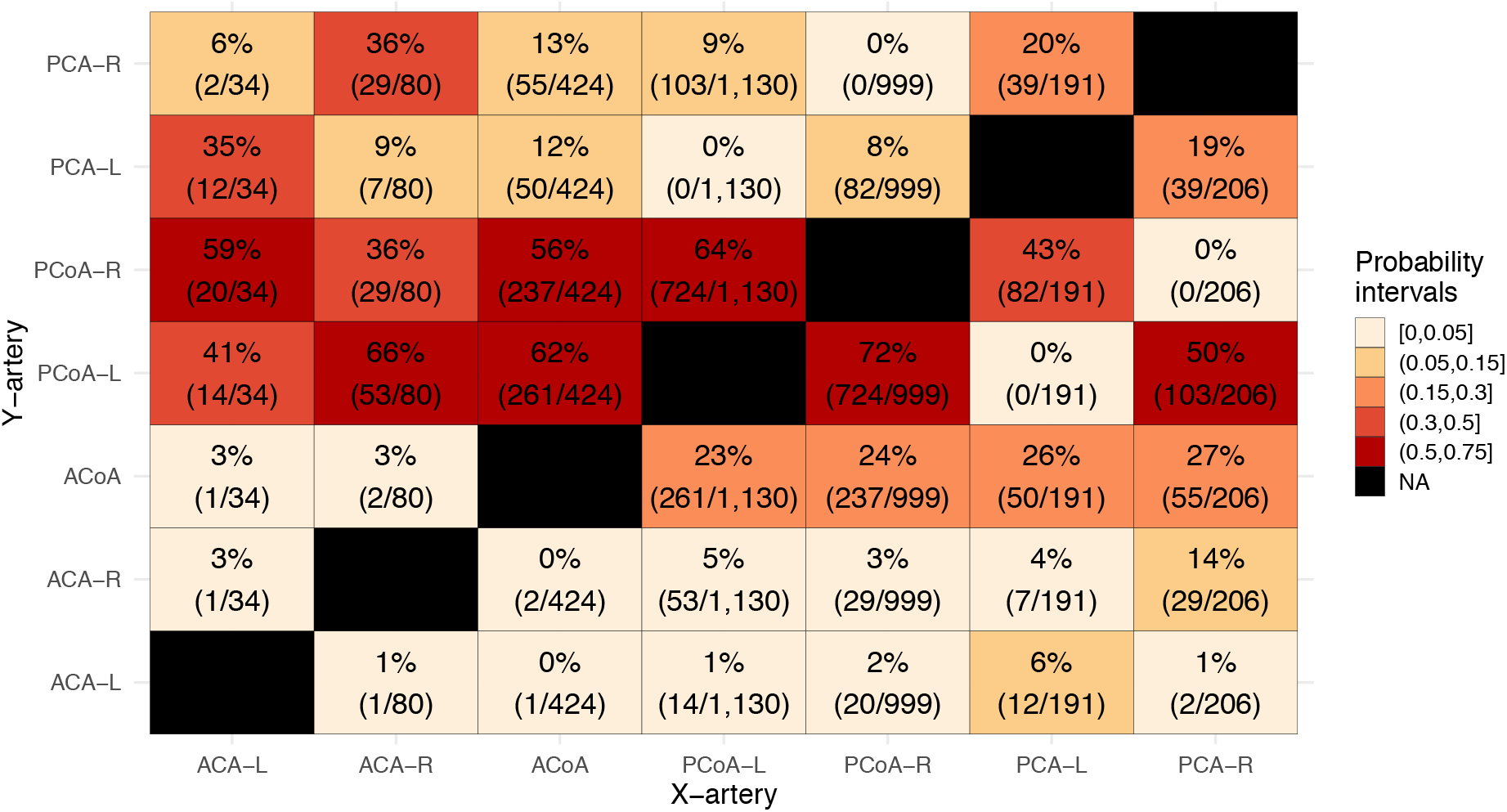
Heatmap with conditional probabilities that artery Y (second axis) is missing given that artery X (first axis) was missing. Numerators and denominators of conditional probability estimates are provided in the brackets, and represent respectively the number of times two segments are missing at the same time (joint probability) and per column the number of times the artery X is missing (independent probability). The common denominator of the joint probabilities and independent probabilities have cancelled. ACA: Anterior cerebral artery. ACoA: Anterior communicating artery. PCoA: Posterior communicating artery. PCA: Posterior cerebral artery. Left and right lateralization are denoted by “L” and “R” respectively. Each successive heatmap interval increases in size with 0.05.

### Tests of conditional independence between CoW variant frequencies and sex, and age, while controlling for the other

The Cochran-Mantel-Haenszel test of conditional independence between sex and CoW variant frequencies while controlling for age (Table 2), resulted in M^2^(22, N = 1,864) = 33.702 with unadjusted P = .0526. This result imply that sex is not significantly associated with the frequency of CoW variants when corrected for mean split age.

The second Cochran-Mantel-Haenszel test (Table 2) tested whether CoW variant frequencies were conditionally independent of being above or below sample mean age while controlling for sex. This test returned M^2^(22, N = 1,864) = 38.849 with unadjusted P = .0147, demonstrating that the mean-split age group was associated with the distribution of CoW variants, when corrected for sex.

### CoW variant frequencies per decade

Figure 5 shows CoW variant frequencies per decade. From this figure, we observed that for each increasing decade the CoW variants that were missing a single artery (Ac, Pcl and Pcr) and the complete variant became less common. We also observed that the composite category of rare CoW variants became more common in later decades. These observations suggest that it is more common in older age to have more missing segments in the CoW.

**Figure 5.**
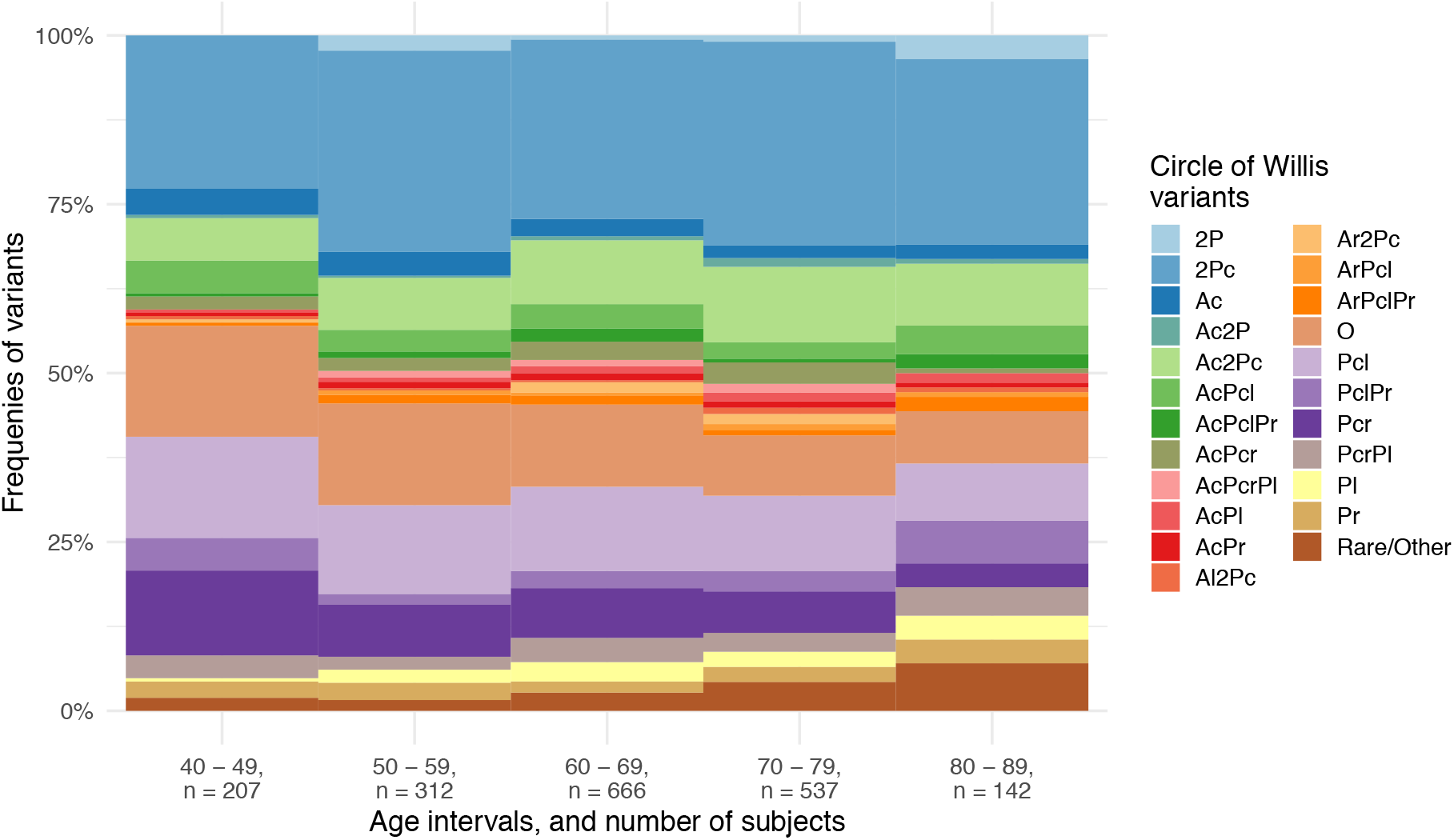
Stacked bar plot of the frequencies of the most common Circle of Willis variants divided into age intervals as decades. Each variant is put together by the missing segments with the following notation: 2P: Missing bilateral posterior cerebral artery. 2Pc: Missing bilateral posterior communicating artery. Ac: Missing anterior communicating artery. Pc: Missing posterior communicating artery. P: Missing proximal posterior cerebral artery. A: Missing proximal anterior cerebral artery. Left and right lateralization are denoted by using “l” or “r” respectively as a suffix for eligible segments. Special cases exempt from the preceding are: O: Complete variant, i.e. no missing segments. Rare/Other: Composite category of other rare variants with one or more missing segments.

### Test of independence between sex and decade age groups

The 5 × 2Chi-squared test was carried out (Table 1) to test for independence between the sex and age as decades group in Figure 5. This test yielded X^+^(4, N = 1,864) = 8.482 with unadjusted P = .075, implying homogeneous distribution of men and women across the five decades. Thus, due to the homogeneity result, Figure 5 is statistically appropriate to interpret equally for both sexes.

### Intra- and inter rater validation

The intra rater validation yielded an accuracy score of 79% (conflicting classification in 21 of 100 cases). Closer inspection showed that only a single artery was mismatched for all 21 variant mismatches (see Supplementary Figure S5). The ACoA was prone to ambiguity with a total of 12 mismatches. ACA and PCA were misclassified as present instead of missing in four and three cases, respectively.

The inter rater validation yielded an accuracy score of 82% (conflicting classification in 18 of 100 cases). Compared to the intra rater validation, the inter rater validation had higher accuracy score, but had on the contrary more severe misclassifications. In other words, the mismatches were not only single artery mismatches. See Supplementary Figure S6 for details on the inter rater validation.

## Discussion

This is, to our knowledge, the largest population-based study on the anatomical variation of the CoW, that included both men and women. The large sample size and recruitment of participants from the general population provide reliable prevalence estimates of the anatomical variation in the CoW for people between 40 to 90 years of age. Main findings were that only 11.9% had a complete textbook CoW variant, while the remaining 88.1% had one or more missing segments in the CoW. In total, we found 47 variants of the CoW, but only five of these variants were very common (i.e. present in > 5%). Further notable findings were that CoW frequencies were associated with age, but not with sex, and that there were patterns of interdependent missing segment patterns across CoW variants.

The excellent agreement in prevalence estimates, with a mean difference of only 1.6%, between our study and a comparable well-powered study (3) has several implications. First, the similarities between a male Chinese population and a Norwegian population suggest that variations in the CoW are similar across populations. A notion consistent with a study on twins that found no genetic effect on the variability of the CoW (6). Second, it supports our finding that gender is not associated with the anatomical variability in the CoW, since the study by Qiu et al. (3) only included men, while our study included both women and men in an approximately equal proportion. Third, since most previous studies have relied on sample sizes of up to a few hundred participants, it is likely, considering the excellent agreement between two studies with a sample size of about 2000, that the disagreement between prevalence estimates in previous studies stem from too small study samples.

We found that CoW frequencies were associated with age, which has been observed in other studies (1,4,23). These studies found, similarly to our study, that the number of missing arteries increased with age. Although the underlying cause of the increase in missing segments with age is not clear, atherosclerosis has been suggested as a possible cause (23). since plaque in an arterial segment might reduce the flow so that the segment is not detected on flow-sensitive TOF MRI. The reduction in cerebral blood flow with age (29) possibly in conjunction to the increase in tortuosity of blood vessels with age (30) could also alter the flow pattern in the CoW such that there is no, or very little flow in some segments, which would also appear as missing segments in the CoW. It is therefore possible that the increased rate of missing segments with age is caused by atherosclerosis or other factors affecting the blood flow in the CoW.

We did not find an association between sex and the frequencies of CoW variants. Previous studies have reported conflicting findings regarding the effect of sex; some find that the complete variant is more prevalent in women (1,4), that specific variations are more common in men or women (9), or that there is no association (23). Differences in methods, sample sizes and statistics, make it difficult to compare our results to the previous findings. However, the large sample size and correction for a possible age bias in our analysis, suggest that the effect of sex on the anatomy of the CoW is at best modest.

Study limitations were as follows. First, the TOF MR technique is sensitive to blood flow, i.e. it is necessary for blood to flow with a sufficient speed to be visible on the TOF images. As such we are only visualising blood flow, not arteries, and some of the missing CoW vessels might well be present, but not visible on the TOF images. This is supported by the higher frequency of the complete CoW variant in dissection studies (31,32), but it is worth noting that dissection studies also show that some sections in the CoW can be completely absent as well (31). We may also argue that it is more clinically relevant to assess the pattern of blood flow in the CoW, not whether the physical arteries are present or not. Second, as seen from the intra- and inter rater validation there were some misclassifications in ambiguous cases of certain arteries. In particular, the ACoA was associated with higher rate of misclassification than other arteries. Some cases of ACA, PCoA and PCA were also mismatched, but not of the same magnitude as ACoA. As such, estimates including ACoA should be considered less accurate. Last, because of the large number of variants found, the precision of frequencies for a given variant should be judged relatively to its number of observations. On the other hand, our study strengths were as follows: (1) a large sample size, (2) a rigorous and reproducible classification scheme, and (3) intra- and inter rater validation indicating similar classification robustness across each rater.

In conclusion, in a large population sample, 47 anatomical variants of the CoW were found, but only 5 variants were commonly encountered. The complete CoW variant was the third most frequent variant present in 11.9% of the sample. Mean-split age was significantly associated with CoW variant frequencies, which could be partially explained by the increased number of missing arteries with increasing age. We also found interdependent missing segment patterns between the ACAs, ACoA, PCoAs, and the PCAs, showing the importance of also including the whole CoW during assessment to retain information about the CoW variants’ collateral ability. Our variant frequencies agreed well with another large-scale MRI study in Chinese men suggesting similar CoW variant frequencies across different populations and that large variability in CoW variant frequency in the literature likely stems from using too small samples. The observed increasing number of missing segments with age suggests that the collateral ability of the CoW may become an increasingly important risk factor for brain health with older age.

## Data Availability

Data generated and consequently analysed during this study are available through The Tromsø Study, currently situated at UiT The Arctic University of Norway.

https://en.uit.no/forskning/forskningsgrupper/gruppe?p_document_id=453582

http://tromsoundersokelsen.uit.no/tromso/

## Acknowledgements

We warmly thank the participants of the Tromsø Study, the administration of the Tromsø Study, the Department of Radiology at the University Hospital North Norway and the MR imaging technologists for their contributions to the study.

## Author contributions

LBH wrote the manuscript, carried out the statistical analysis and discussion, created the figures, created the classification scheme and program, readied the data to a useable form, and classified the data. AKH revised the manuscript and contributed to study design and planning. LHJ helped in collecting and organizing the data, and contributed to study design and manuscript preparation. EBM contributed to study planning, study design and manuscript preparation. DR helped revising the manuscript, and contributed to the classification scheme and programming. TRV was the principal investigator supervising the study and contributed to most aspects of this study.

## Data availability

Data generated and consequently analysed during this study are available through The Tromsø Study, situated at UiT The Arctic University of Norway.

## Supplementary information

Supplementary information consisting of a pdf file and a xlsx file are available.

